# Potential severity, mitigation, and control of Omicron waves depending on pre-existing immunity and immune evasion

**DOI:** 10.1101/2021.12.15.21267884

**Authors:** Ferenc A. Bartha, Péter Boldog, Tamás Tekeli, Zsolt Vizi, Attila Dénes, Gergely Röst

## Abstract

We assess the potential consequences of the upcoming SARS-CoV-2 waves caused by the Omicron variant. Our results suggest that even in those regions where the Delta variant is controlled at the moment by a combination of non-pharmaceutical interventions and population immunity, a significant Omicron wave can be expected. We stratify the population according to prior immunity status, and characterize the possible outbreaks depending on the population level of pre-existing immunity and the immune evasion capability of Omicron. We point out that two countries having similar effective reproduction numbers for the Delta variant can experience very different Omicron waves in terms of peak time, peak size and total number of infections among the high risk population.

## 1. Introduction

At the end of November 2021, according to the World Health Organization, the B.1.617.2 Delta variant accounted for 99% of COVID-19 cases around the world [1]. Yet in recent weeks, we have observed the rise of the B.1.1.529 lineage, designated as the Omicron variant. This variant was first reported to the WHO on 24^th^ November, and as of 9^th^ December, Omicron sequences have been found already in 63 countries [2], in spite of travel restrictions. Omicron transmission dynamics can be estimated from sharp turning points in epidemiological trends, widespread genome sequencing or by S–gene target failure (SGTF), which is a proxy for Omicron [6]. Omicron has out-competed Delta in South Africa in a short time [3], and rapid growth of cases has been observed in the United Kingdom [4] and Denmark [5].

There is accumulating evidence of high transmissibility and immune-evasion capability of the Omicron variant ([4, 6, 7]). Ongoing neutralization studies indicate a significant drop in vaccine efficacy [8], and increased frequency of reinfection has been reported in South Africa [7].

There is an urgent need to estimate the potential impact of this variant. Modelling for the United Kingdom has been posted on 11^th^ December [4]. However, since countries have different levels of pre-existing immunity either from vaccination or from previous infections, and different non-pharmaceutical measures are in place, we can expect that countries will be affected differently. Our goal here is to provide a quick assessment of this threat, considering these country-specific factors.

## 2. Methods

### 2.1. Potential control of Delta and Omicron

The effective reproduction number corresponding to the Delta variant (denoted by 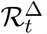) at a given time characterizes the current transmission of the infection in a population. It can be obtained by the correction of the basic reproduction number 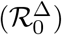 with the mitigating effect of the actual non-pharmaceutical interventions (NPI) and the population immunity level (*p*) in reducing transmission. That is, 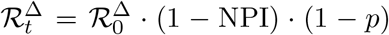. Similarly, if the Omicron is already present in the same population, its effective reproduction number is 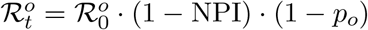, where 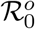 is the basic reproduction number of the Omicron variant, and *p*_*o*_ is the population immunity against this variant.

We introduce parameter *e* for the extent of immune evasion, expressing that immunity *p* to prior variants is reduced by *e* · *p* with respect to Omicron, that is *p*_*o*_ = *p* · (1 − *e*). Let *p*^*SA*^ be the population immunity in South Africa, and let 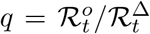 at the time of the emergence of Omicron. Then, we have the relation

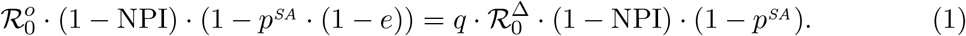

In a country with a combination of pre-existing immunity *p* and NPIs, to achieve 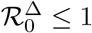 and 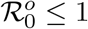, one needs

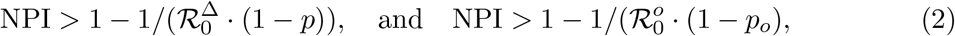

respectively.

### 2.2. Transmission dynamics of Omicron

To estimate the potential of an Omicron wave, we employ a compartmental model of disease dynamics, which is monitoring the temporal change of the number of infected individuals in the population, separately with and without pre-existing immunity. The technical details of the model and the numerical code are described in the Supplement. Most parameters are estimated from the literature, while *p* and *e* are varied in a feasible interval. The differential equations are solved for many combinations of *p* and *e*, thus we can assess the total epidemic size, the peak size and the peak time of the Omicron wave for a range of scenarios.

### 2.3. Parametrization

Current estimates from South-Africa and United Kingdom indicate that in those countries 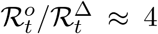 [11, 12, 20], which means that an Omicron-case generates four times more cases than a Delta-case. As baseline parameters, we choose 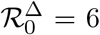 [9, 10], and for South Africa at the time of the emergence of Omicron we assume 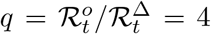, and *p*^*SA*^ = 0.85 [13], but these key parameters can easily be varied to explore the sensitivity of the outcome. The choice of *p*^*SA*^ is consistent with the observed 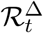 in South Africa in a period before Omicron [11]. Previous experience in European countries shows that strict lockdowns can achieve an 82% drop in the effective reproduction number [14], hence we consider NPI ≤ 0.82. For the compartmental model, following [4], the average incubation period is assumed to be 2.5 days, and the average infectious period is assumed to be 5 days, each following a gamma distribution.

## 3. Results

Since the NPIs affect the transmission of both variants in the same way, from 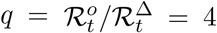, we have 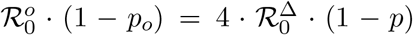. Such a four-fold advantage of Omicron can emerge either from inherently higher transmissibility or a larger susceptible pool, when immunity obtained by vaccination or prior infection by other variants does not protect against the new variant as effectively. There is an obvious trade-off relationship between these two factors determining the transmission fitness: the higher the transmissibility, the lower the immune evasion must be to maintain the four-fold ratio of the effective reproduction numbers.

With our baseline parameters, from (1), we obtain the relation

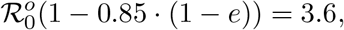

showing that if immune evasion is significant, then 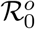 must be more moderate. In particular, 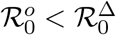 whenever *e* > 0.53.

Consider a country with population immunity *p*, where Delta is controlled, then by (2), NPI > 1 − 1*/*(6 · (1 − *p*)). The necessary NPIs to control Omicron can also be found from (1) and (2) as

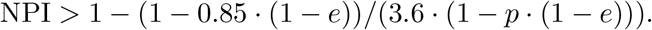

The necessary NPIs are plotted in Fig. 1) as a function of *p* for various values of *e*. As we can see, the NPIs that are sufficient against Delta are not enough to stop the spreading of Omicron, for any considered combination of *p* and *e*. Since Omicron containment requires very stringent NPIs, the invasion of this variant is likely to result in widespread infection.

**Fig. 1.**
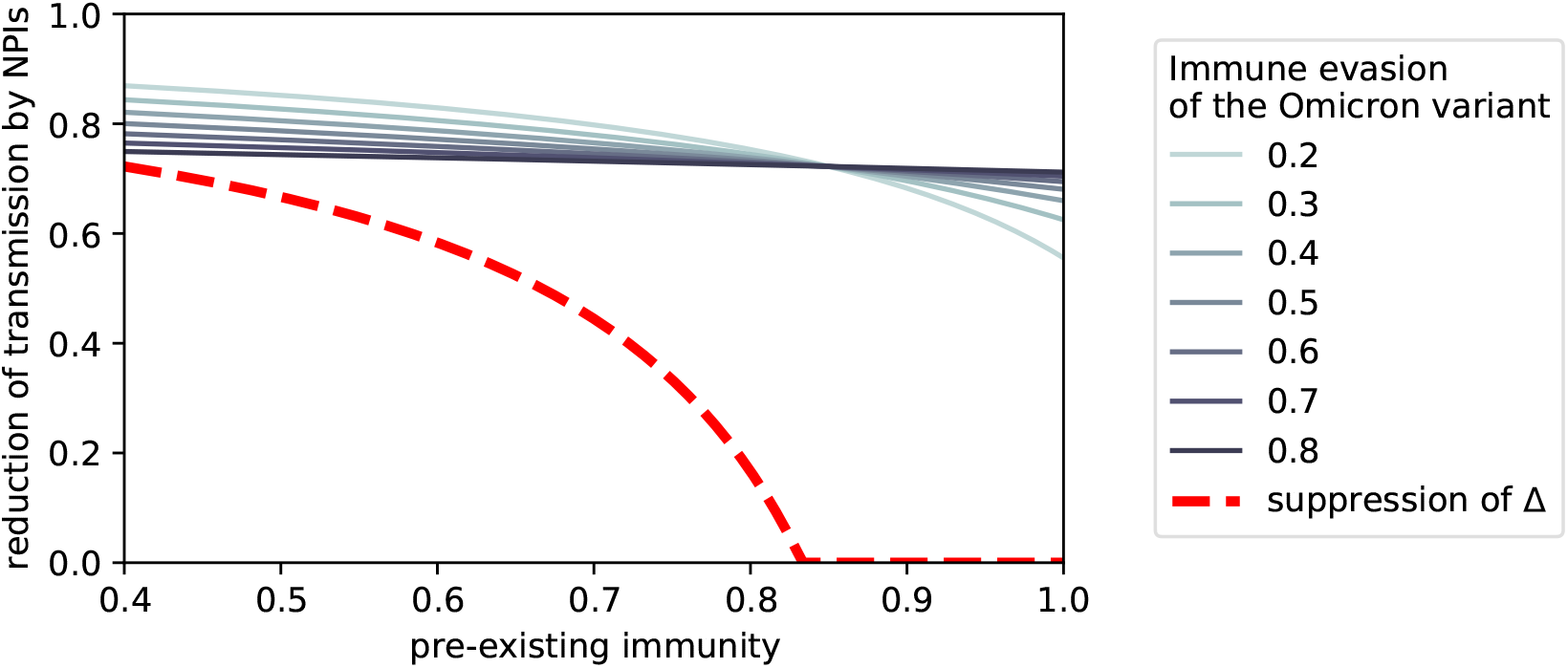
The necessary level of non-pharmaceutical interventions (NPI) to control the Omicron and Delta (dashed) variants as a function of pre-existing immunity

To estimate the severity of future Omicron waves in countries where Delta is under control, we solve our transmission model for a range of parameters (*p, e*), and when population immunity is below the Delta herd immunity threshold, we employ the necessary NPIs (the red dashed curve in Fig. 1) to achieve 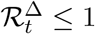. The heatmap in Fig. 2) shows the fraction of the population infected during the Omicron wave with respect to the parameters (*p, e*) space. One can see that the only scenarios not to have a significant outbreak are the following: the population has extremely high immunity and Omicron is not very immune evasive (bottom right corner); or the competitive advantage of Omicron emanates from high immune evasion rather than inherent transmissibility and at the same lockdown-like NPIs are in place (top left corner): none of which are plausible. We chose examples *a, b, c, d* from the highlighted rectangle of a feasible parameter region for more detailed investigation, but in our published code [21] we provide an interactive tool that makes exploring other scenarios very easy for the reader.

**Fig. 2.**
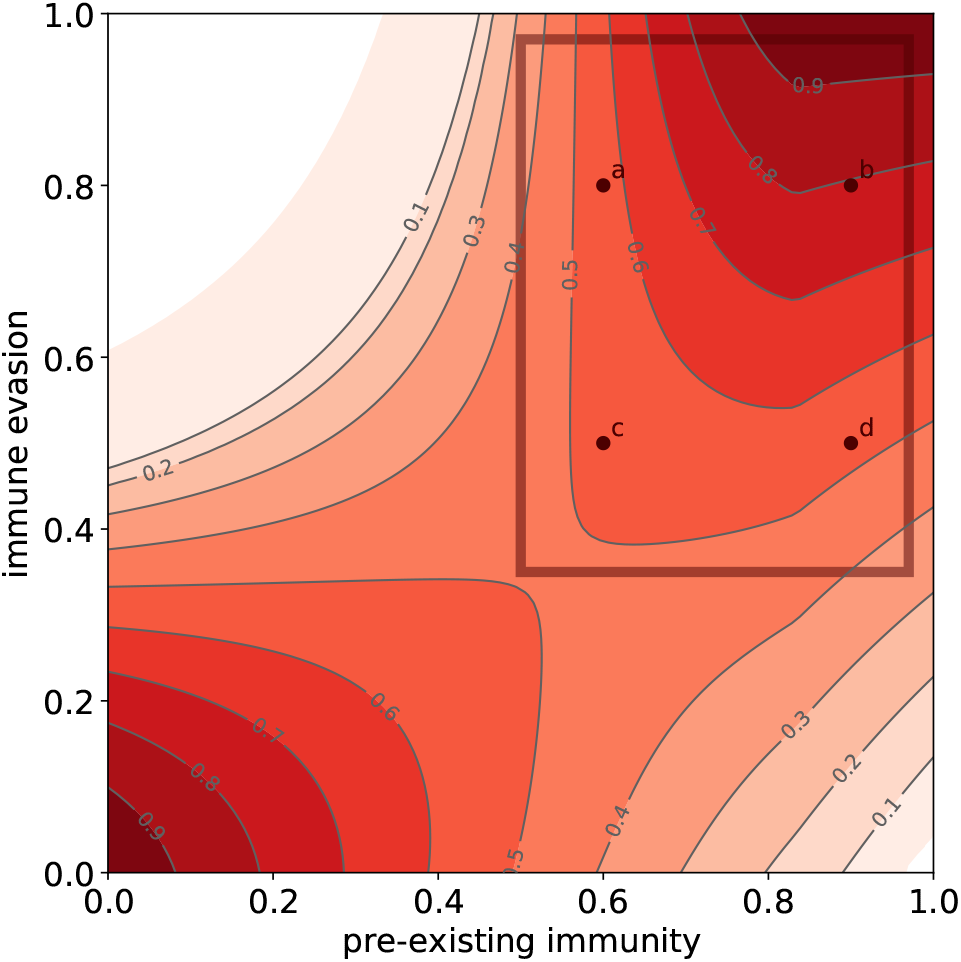
Heatmap of the total number of infections (as fraction of the population) during Omicron wave, depending on pre-existing immunity and Omicron’s immune evasion, assuming that no further mitigation measures will be implemented. We select examples from a feasible parameter region highlighted by a rectangle.

The time course of the Omicron outbreak in the four selected scenarios are depicted in Fig. 3. The dark red colour represents infected population without prior immunity, while the pink colour represents infected population having been vaccinated or previously infected. In the insets, the cumulative infected fraction is shown, and the grey colour represents the population with prior immunity, while the lighter shade is the portion becoming available for Omicron-infections due to immune evasion.

**Fig. 3.**
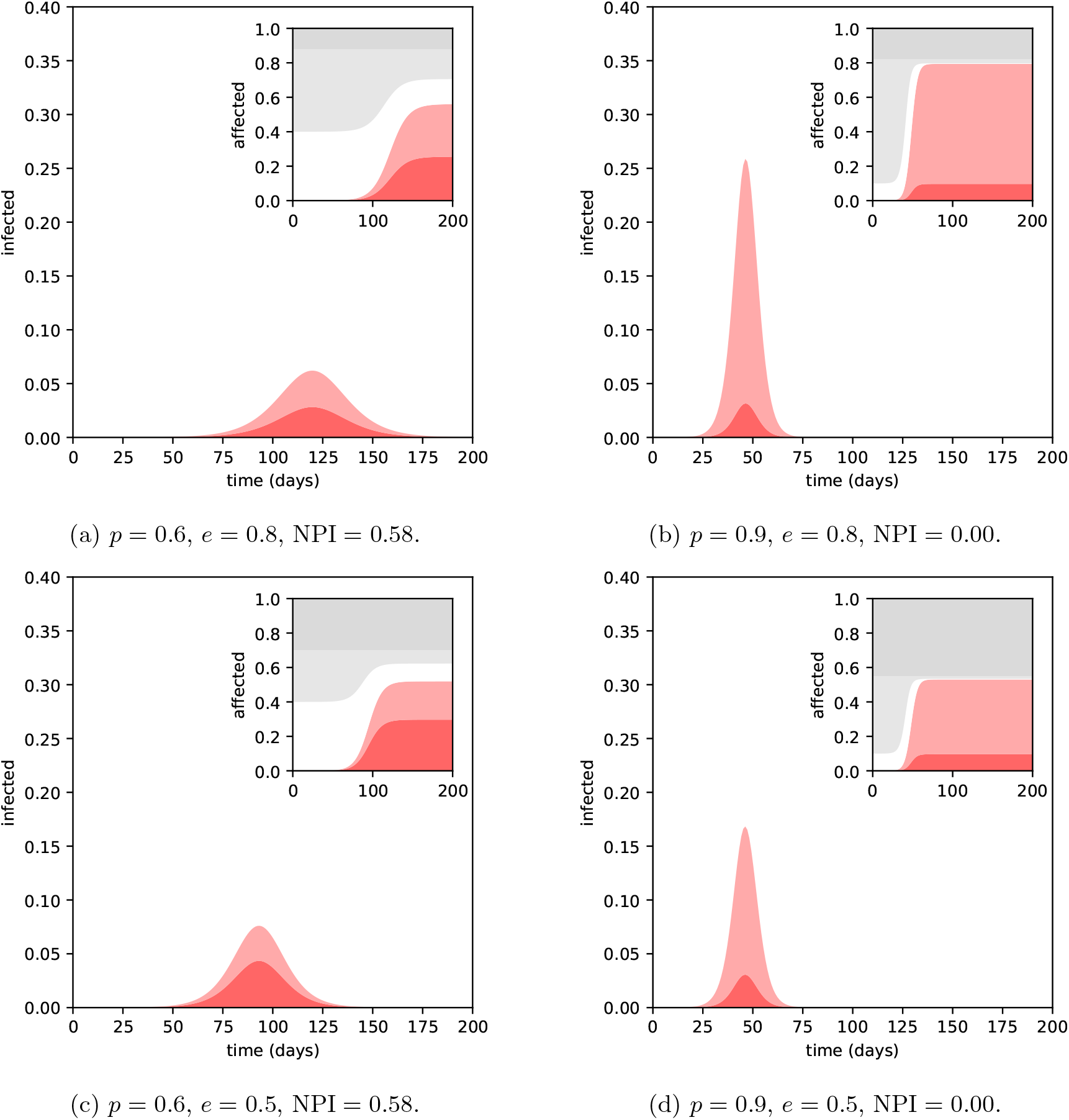
Epidemic curves of the Omicron wave under various assumptions on the pre-existing immunity and the variant’s immune evasion, without any additional measures.

Potential Omicron waves may have very different characteristics. In countries with very high population immunity, where Delta is contained with very mild or without NPIs, one can expect an extremely rapid increase of cases and a high peak in a matter of weeks, especially if Omicron is highly immune evasive (scenarios *b* and *d* in Fig. 3). If population immunity is moderate and strong NPIs are being employed to contain Delta, then the Omicron wave is more flattened, as long as the NPIs remain in place (scenarios *a* and *c* in Fig. 3). In this case, as opposed to the high immunity scenarios, the peak is lower if immune evasion is higher.

Besides the sheer number of infections, to assess the severity of the outbreaks, it is very important to take into consideration the prior immune status of the population getting infected. For example, despite in *b* and *d* the peak is much higher than in *a* and *c*, if Omicron infections of those with prior immunity turn out to be overwhelmingly mild, than the severity is much better reflected by the number of infected without prior immunity, that is the red curves without the pink part in Fig. 3). In this measure, the *a* and *c* are more severe.

For scenarios *b* and *d*, reintroducing measures with NPI = 0.4 can reduce the peak size roughly to half, and delaying the peak by a month, as shown in Fig. 4).

**Fig. 4.**
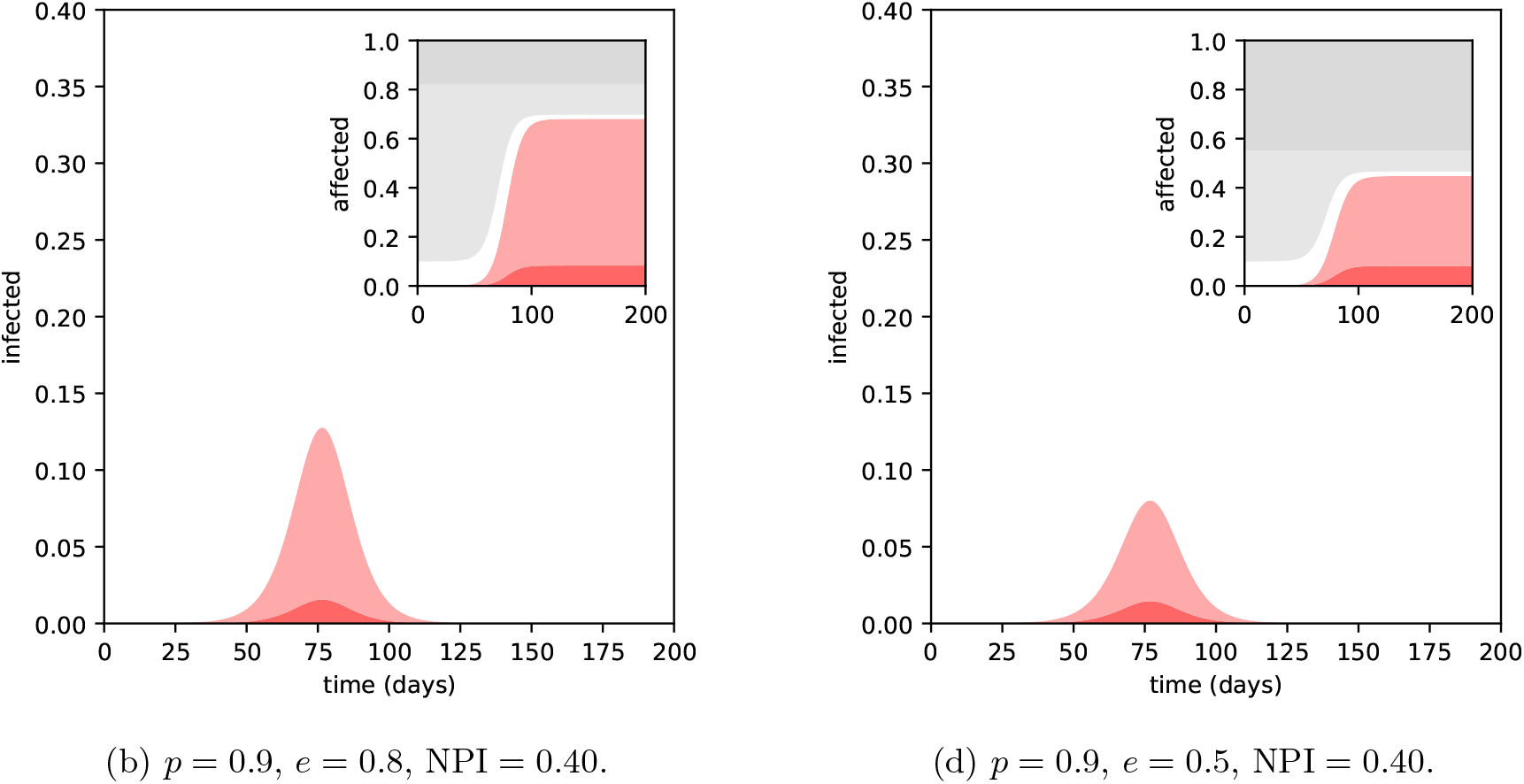
Effect on the Omicron wave: introducing moderate NPIs in countries with high immunity against the Delta variant. Scenarios (3b) and (3d) from Fig. 3).

## 4. Discussion

The Omicron variant is spreading across the globe with unprecedented speed. Hence there is great urgency to assess its potential consequences, despite all uncertainties about its epidemiological parameters. Our calculations suggest that the current measures, in combination with population immunity, that might be sufficient against the Delta variant, will not be able to suppress Omicron. Hence widespread Omicron transmission is expected. However, the impact of the Omicron wave on various countries can be very different depending on the level of pre-existing population immunity, and the immune evasion property of Omicron. The severity of the disease caused by Omicron is unclear, however previous immunity is expected to reduce the risk of severe outcome [4]. To further improve the assessment, we constructed a transmission dynamics model that tracks the emerging infections in the groups with and without pre-existing immunity separately. This allowed us to investigate a range of scenarios, and we found that in countries with high immunity but mild NPIs a very sharp epidemic curve is expected with a high peak, but a smaller fraction of infections are from the high risk population. On the other hand, countries with moderate immunity that employ stricter NPIs at the moment to fight Delta can have a flatter Omicron curve, but with more infections from the high risk group. Reintroducing further NPIs might be needed to mitigate the Omicron waves, and that would also buy some time to expedite booster roll-outs.

## Data Availability

The present study did not produce any new data.

## Acknowledgement

The authors were supported by NKFIH and ITM (FK 138924, FK 124016, PD 128363, KKP 129877, 2020-2.1.1-ED-2020-00003, TUDFO/47138-1/2019-ITM, UNKP-21-5). F.B. was also supported by the Bolyai Scholarship of the Hungarian Academy of Sciences.

## Author’s contributions

Conceptualization and methodology, G.R.; codes and computations F.B., P.B., Zs.V.; data collection and analysis, A.D., T.T.; writing and editing, F.B., A.D., T.T., G.R.; visualization, P.B., F.B., G.R., Zs.V.

## Supplementary material

### S1. Trade-off between inherent transmissibility and immune evasion capability

The contour formula (1) results in a trade-off between immune evasion and the spreading capability of the Omicron variant when all other parameters are kept fixed. In Fig. 5), we demonstrate this phenomenon fixing 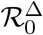 and *q* to their baseline values and allowing uncertainty of the pre-existing immunity (*p*^*SA*^) in South Africa.

**Fig. 5.**
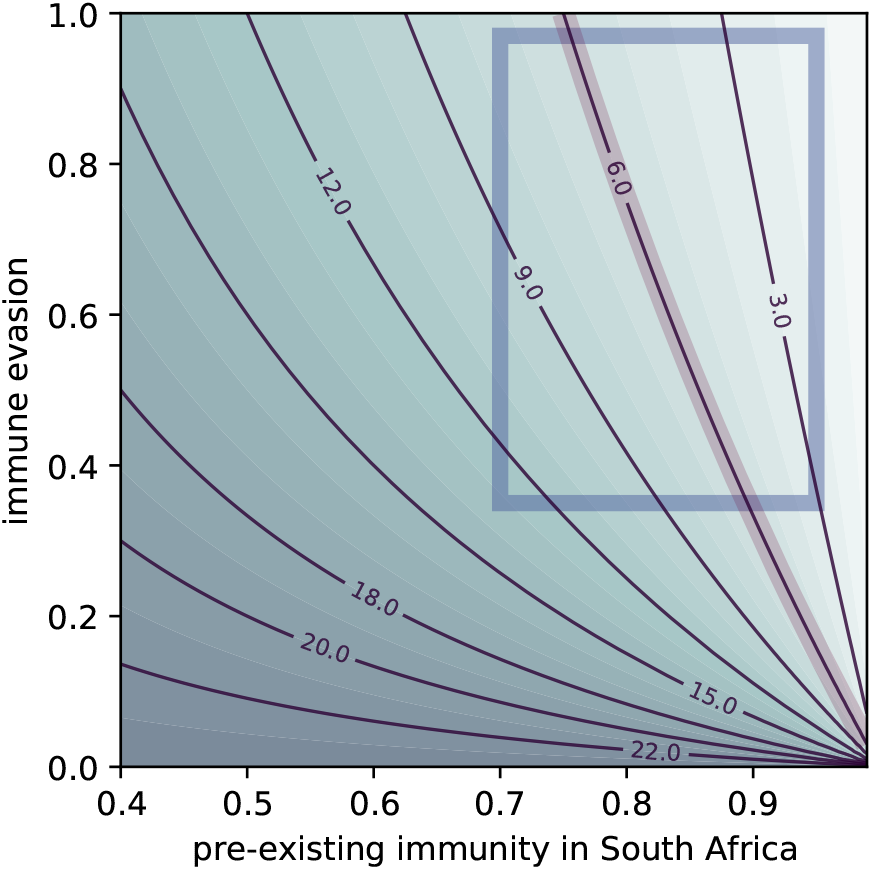
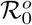 contours visualizing the trade-off between transmissibility (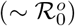) and immune evasion (e) of the Omicron variant. The rectangle marks a feasible parameter region for South Africa, the highlighted contour agrees with the transmissibility of Delta.

Note that the scenarios with 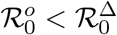 on Fig. 5) are to the right of the contour with special focus. Clearly, even when considering only *p*^*SA*^ ≈ 0.85, the uncertainty of 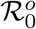 is substantial as potential values lie within a range of [1.5, 24], where 24 = *q* · 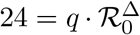 is the theoretical upper limit when there is no immune evasion whatsoever. If we restrict our attention to a more feasible immune evasion region, similar to what was highlighted on the *y*-axis of Fig. 2), we may bound our estimate to 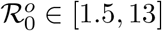.

### S2. Transmission dynamics model

The transmission dynamics is based on an *SE*_2_*I*_4_*R* model, generalized from *SEIR* by assuming gamma-distributed incubation and infectious periods, using the Erlang parameters *n* = 2 and *m* = 4. The sixteen compartments 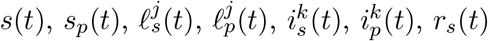 and *r*_*p*_(*t*) – with *j* = 1, 2; *k* = 1, 2, 3, 4 – represent fraction of the population being in different disease and immunity status. The compartment *s* designates population level susceptibility, without any pre-existing immunity to any of the variants, while *s*_*p*_ stands for susceptibles to Omicron despite of being immune to earlier variants due to a past infection or vaccination. Susceptibles (either from *s* or *s*_*p*_) can get infected upon adequate contact with an infectious person (from one of 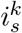 or 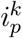) with transmission rate *β*, and moved to the corresponding first latent compartment. For both pathways **i** = *s, p*, the latent class comprises the two stages 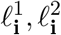. The length of the latent period is *α*^−1^, thus, one moves with the rate 2*α* from the first latent stage to the second and thereon to the first infectious compartment, alike. The model features four infectious stages stretching over the infectious period *γ*^−1^, again, for both pathways. Individuals transition from one infected stage to the next one, as well as removed from the fourth with the rate 4*γ*. This linear chain ensures that the infectious period is gamma distributed. We do not model the clinical outcome of disease progression and all non-infectious individuals eventually enter an *r* class. Note that this does not pose any restrictions on assessing the severity of the Omicron wave as said severity is in direct correlation with the transmission dynamics of the epidemics captured by model time series and the corresponding peak and final size.

The above considerations are summarized in the compartmental ODE system

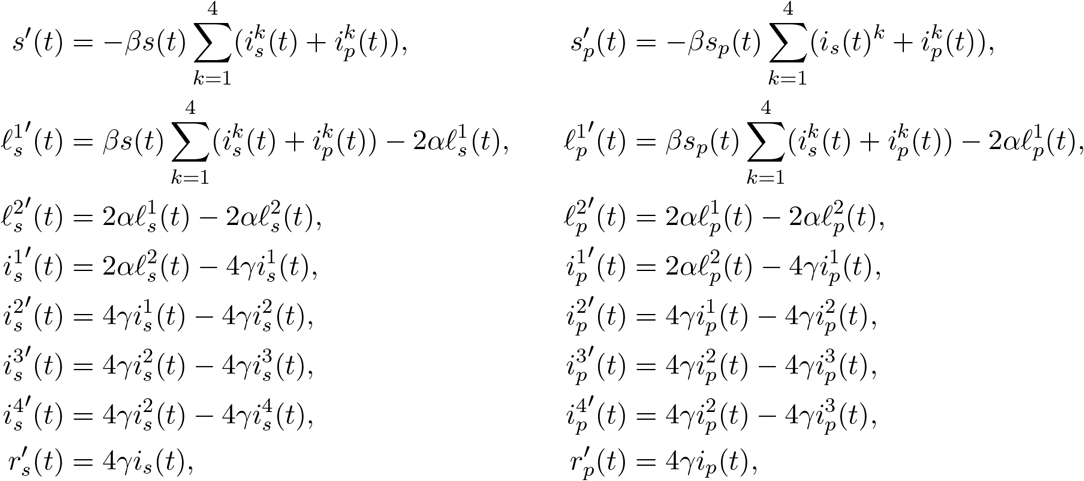

with the transmission rate *β* = *R*_0_*γ*(1 − NPI). Here, the level of non-pharmaceutical interventions NPI(*t*) is assumed to be constant in the timeframe of each numerical simulation. Finally, the computations were carried out using the initial values *s*(0) = 1 − *p, s*_*p*_(0) = *e* · *p*, and a small amount of initial Omicron-infected.

### S3. Supplementary source codes

The software used in our analysis and for generating all figures is available on Github [21] with an option for direct experiments in Google Colab. We implemented our solution as a Jupyter notebook with Python kernel using the standard libraries for computation (`scipy`, `numpy`) and visualization (`matplotlib`).

The code is structured as follows. First, global parameters are defined such as 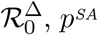, and *q* enabling effortless sensitivity analysis of the results. Then, the methodological core is set up to compute 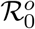, suppressing NPIs in various scenarios, and to carry out numerical simulations of the ODE model. Finally, the code snippets for visualization are enabled and a multitude of figures are produced, many of which are equipped with an interactive interface to ease the exploration of the parameter space.

### S4. Invasion reproduction number of Omicron

The effective reproduction number at the time of introduction into a population is shown in Fig. 6. The values in the figure are in line with the empirically estimated value of 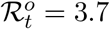 for the UK [4].

**Fig. 6.**
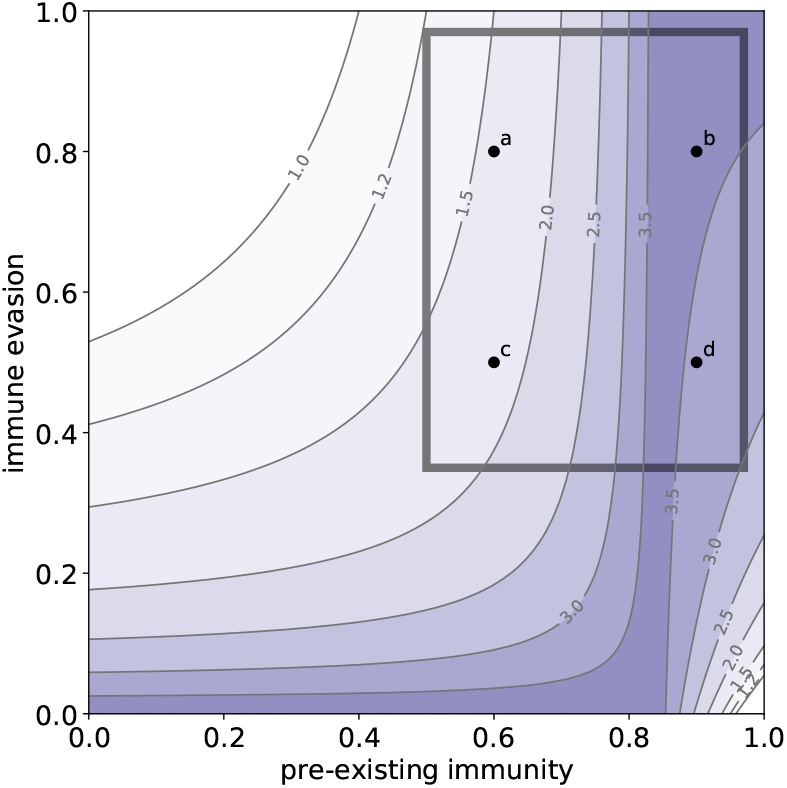
Reproduction number of Omicron when introduced into a population where Delta is controlled.

### S5. Severity assessment

The overall severity of an Omicron wave might be assessed by the number of infected individuals who had no prior immunity, as they are at higher risk of severe disease outcome. Fig. 7 shows the comparison of heatmaps. A striking difference is that while the peak size of all infected is the highest when pre-existing immunity and immune evasion are both very high, meanwhile this is a relatively favorable situation in terms of peak size of the infected without prior immunity. A similar comparison can be made between the total number of infections (Fig. 2) and counting only those without prior immunity Fig. 8.

**Fig. 7.**
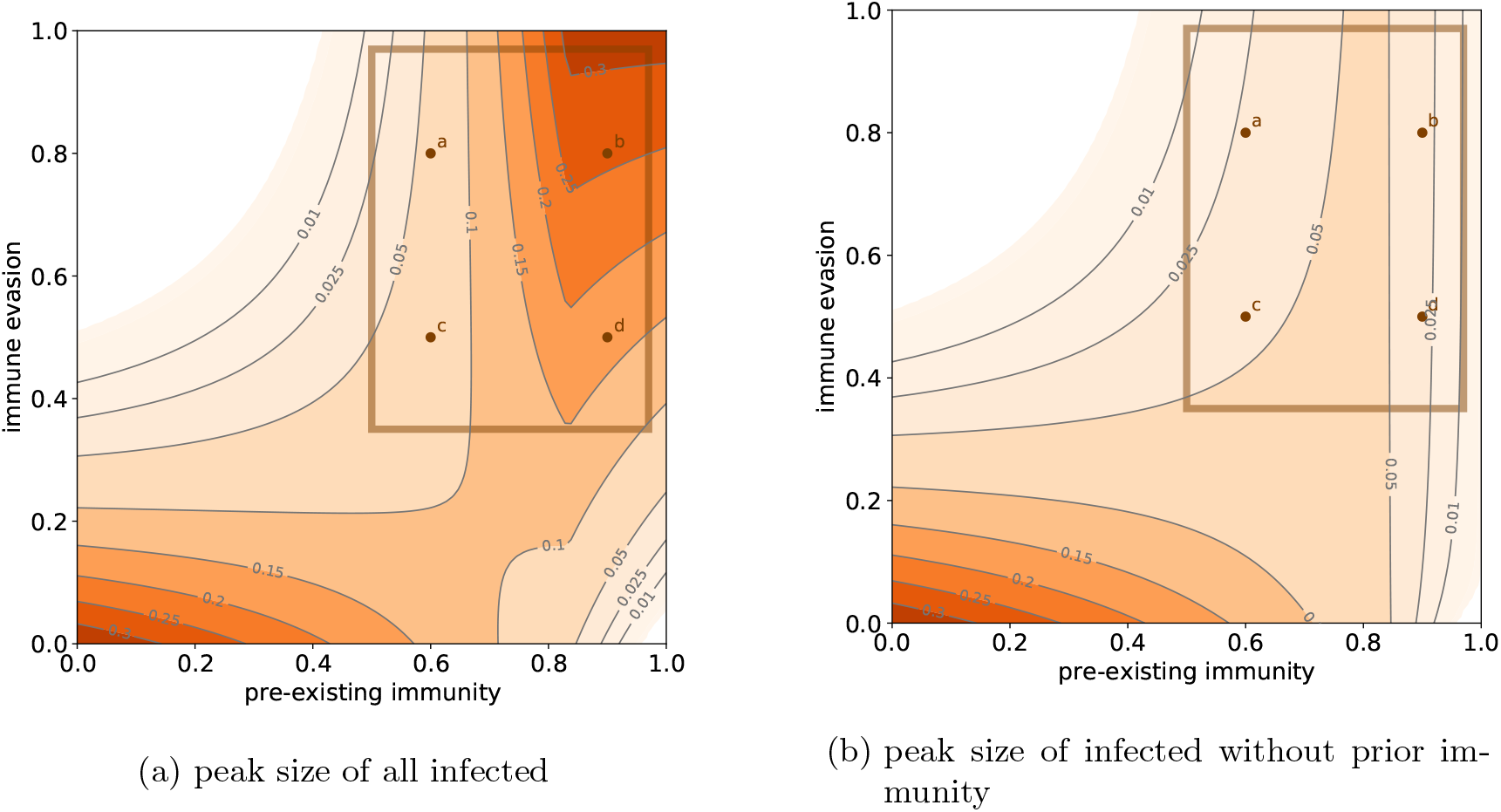
Peak size of the Omicron wave including and excluding the population with pre-existing immunity.

**Fig. 8.**
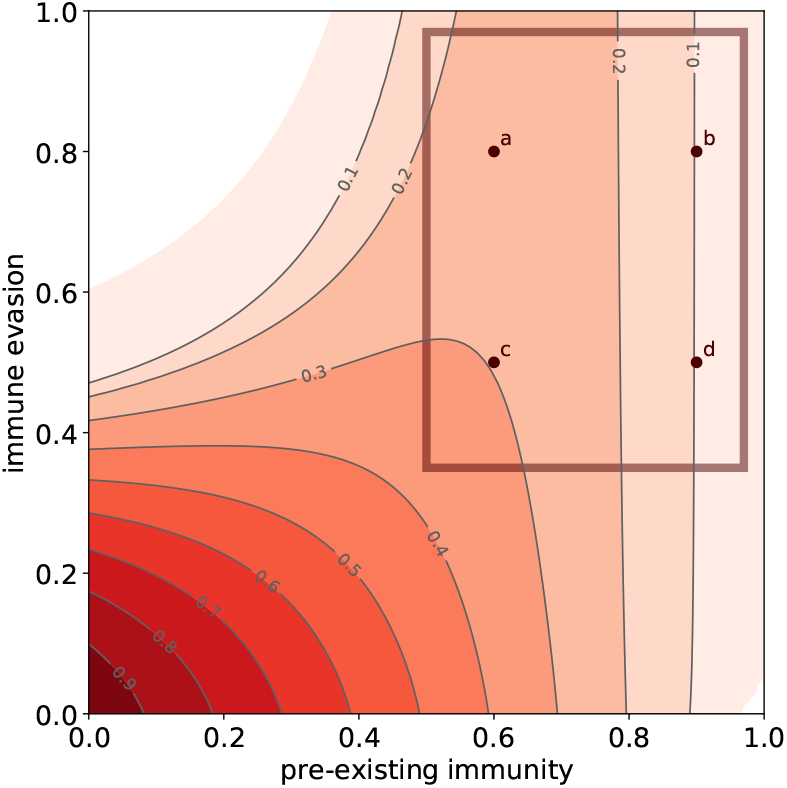
Fraction of the infected population during the Omicron wave counting only those who have no pre-existing immunity.

### S6. Sensitivity to parameters

Our publicly available code [21] make it easy for anyone to explore the sensitivity of the outcomes to the key parameters. A higher value *p*^*SA*^ would mean that the Omicron is less transmissible hence more controllable by NPIs, see Fig. 9. Varying 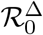 around 6 does not change the main features of the figures. Decreasing *q* makes the epidemic curves flatter, however for *q* = 3 the invasion reproduction number of Omicron is becoming already too small compared to the observations from UK [4].

**Fig. 9.**
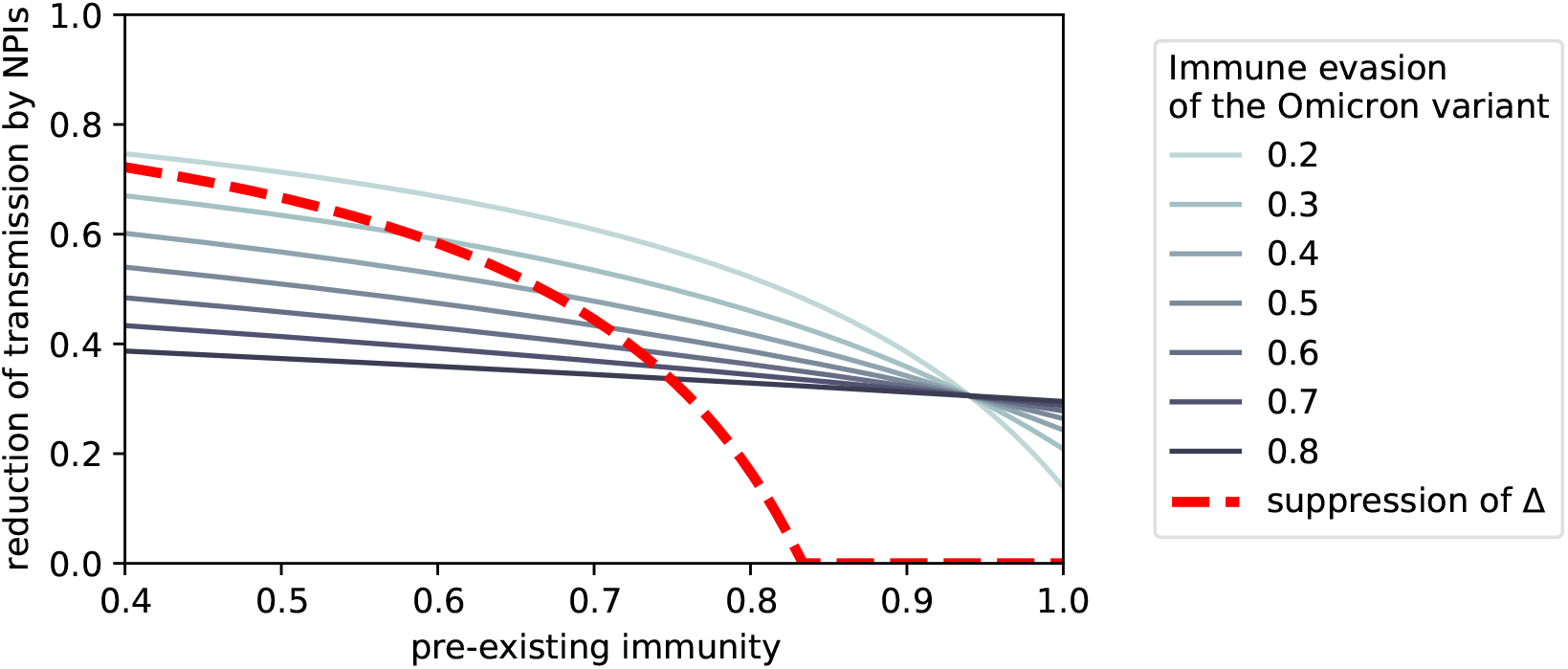
The necessary level of non-pharmaceutical interventions to control the Omicron and the Delta variants as a function of pre-existing immunity, when *p*^*SA*^ = 0.94.

